# Retrospective Evaluation of Hematological Indicators in Predicting CKD Progression

**DOI:** 10.1101/2025.05.26.25328364

**Authors:** Collince Odiwuor Ogolla, Lucy W. Karani, Stanslaus Musyoki, Phidelis Maruti

## Abstract

**Background:** The chronic kidney disease progression is affected by numerous factors, such as hematological abnormalities. Once reliable hematological prognostic factors are identified, early risk stratification and management of patients can be improved.

**Objective:** To retrospectively study whether hematological parameters are correlated with chronic kidney disease progression among patients at Kisii Teaching and Referral Hospital.

**Methods:** We reviewed records of 120 CKD patients who had baseline and follow-up data from April 2024 to April 2025. CKD stages were classified according to KDIGO 2012 guidelines. Progression was considered if there was a decline of at least one CKD stage or initiation of dialysis during follow-up. An array of hematological markers was studied: hemoglobin (Hb), hematocrit (Hct), red cell distribution width (RDW), mean corpuscular volume (MCV), mean corpuscular hemoglobin (MCH), mean corpuscular hemoglobin concentration (MCHC), white blood cells (WBC), neutrophils, lymphocytes, neutrophil-to-lymphocyte ratio (NLR), and platelets. Group comparisons were done using the t-test and ANOVA, and independent predictors were identified by multivariate logistic regression.

**Results:** Some 48.3% of patients progressed to CKD. With increased stages of CKD, Hb, Hct, and lymphocyte counts dropped significantly, while RDW and NLR increased significantly (p < 0.001). Progressors had significantly lower Hb, Hct, MCV, MCH, and lymphocyte counts but higher RDW and NLR than non-progressors (all with p < 0.05). The results of the logistic regression showed that lower Hb, higher RDW, higher NLR, and lower lymphocyte counts were independent predictors of progression (Hb: aOR=0.57, p<0.001; RDW: aOR=1.51, p=0.005); (NLR: aOR=1.89, p=0.002); (lymphocytes: aOR=0.46, p=0.048).

**Conclusion:** Hematological markers, mainly Hb, RDW, NLR, and lymphocyte count, remain important prognostic markers for CKD progression and may be incorporated into routine clinical monitoring with a view to intervening early.

## Introduction

Chronic kidney disease (CKD) is considered a global public health problem that involves a progressive loss in kidney function leading to end-stage renal disease (ESRD), increased morbidity, and mortality [1]. Early identification of patients at high risk of CKD progression is crucial to allow for timely intervention that can delay the bad outcome [2]. However, based on the progress in our comprehension of CKD pathophysiology, cost-effective prognostic markers are still very few, especially in settings where limited resources are available.

Hematological abnormalities may be common in CKD due to impaired erythropoiesis, chronic inflammation, and metabolic disturbances [3]. Anemia being a recognized complication exacerbates kidney function decline; it further enhances cardiovascular disease and lowers life quality [4]. Several hematological parameters contemporary evidence suggests, including red cell distribution width (RDW), neutrophil-to-lymphocyte ratio (NLR), and lymphocyte count, may also reflect inflammation and oxidative stress-the two principal factors involved in CKD progression [5,6].

Several studies have found these hematological indices altered in association with adverse renal outcomes and death [7]. In African populations, however, data remain scarce and validation of findings based on other clinical settings remains to be done. Kisii Teaching and Referral Hospital is in a position to test for the prognostic value of hematological tests that are routinely performed in the care of its large number of CKD patients.

This retrospective study aims to assess the linkage between hematological indicators and CKD progression among patients attending Kisii Teaching and Referral Hospital. We hypothesize that specific hematological parameters can predict CKD progression, potentially guiding risk stratification and management in clinical practice.

## METHODS

### Study Design and Setting

This was a retrospective study conducted at the Kisii Teaching and Referral Hospital (KTRH), a regional referral center in Kisii County, Kenya.

### Study Population

This study looked at CKD patients seeking healthcare services at KTRH. A total of 120 were included in the analysis, whose medical records were retrospectively analyzed. Inclusion criteria: adults aged ≥18 years, diagnosed with CKD based on KDIGO 2012 criteria, with complete hematological and renal function data at baseline and at least 12 months of follow-up. Exclusion criteria: patients with acute kidney injury or unclear diagnosis, those with hematologic malignancies or on active chemotherapy, missing laboratory records, or females who are pregnant.

### Data Collection

Data were retrospectively extracted from electronic and paper-based patient records between April 2024 and April 2025. The data extraction commenced on May 5th and ended on May 10th, 2025, using a standardized data abstraction form. The variables extracted included: demographic details: age, sex; clinical details: CKD stage at baseline (by eGFR), dialysis initiation; hematological parameters: hemoglobin (Hb), hematocrit (Hct), red cell distribution width (RDW), mean corpuscular volume (MCV), mean corpuscular hemoglobin (MCH), mean corpuscular hemoglobin concentration (MCHC), total white blood cells (WBC), neutrophils, lymphocytes, neutrophil-to-lymphocyte ratio (NLR), and platelets. CKD staging was based upon calculating the eGFR using KDIGO 2012 classification: Stage 2: eGFR 60-89 mL/min/1.73 m^2^, Stage 3a: 45-59, Stage 3b: 30-44, Stage 4: 15-29, Stage 5: <15 or on dialysis. Progression was defined as a decline by at least one CKD stage from baseline or initiation of dialysis during the follow-up period.

### Statistical Analysis

Data entry was performed in Microsoft Excel and analysis using IBM SPSS Statistics version 26. Continuous variables were presented as mean ± SD, comparison among the groups of the progressors and non-progressors: Independent samples t-test for variables with normal distribution, One-way ANOVA to examine hematological trend along CKD stages, Chi-square test for categorical variables. Multivariate logistic regression analysis was used to identify independent hematological predictors of CKD progression with a ORs and their respective 95% CIs reported. p-values of less than 0.05 were considered statistical significance.

### Ethical Considerations

Institutional ethical clearance was obtained from Kisii Teaching and Referral hospital ethical review committee (ISERC/KTRH/077/25) and research permit obtained from National Commission for Science and Technology (NACOSTI) - NACOSTI/P/25/417549. Access permission was obtained from KTRH, authors had access to information that could identify individual participants during or after data collection. All patient data were anonymized by removing identifying information, and access to the records was restricted to authorized study personnel to ensure confidentiality throughout the research process.

## Results

### Baseline Characteristics

A total of 120 patients with CKD were enrolled in the study. The mean age of the subjects was 56.4 ± 14.1 years, with 53.3% (n = 64) males. The baseline CKD stages were categorized as per KDIGO 2012 classification as in table 1:

**Table 1.** Baseline Characteristics and CKD Staging (N = 120)

| Variable | Category/Value | Frequency (%) / Mean ± SD |
| --- | --- | --- |
| Age (years) | – | $56.4 \pm 14.1$ |
| Sex | Male | 64 (53.3%) |
|  | Female | 56 (46.7%) |
| CKD Stage at Baseline | Stage 2 (eGFR 60–89) | 12 (10.0%) |
|  | Stage 3a (eGFR 45–59) | 25 (20.8%) |
|  | Stage 3b (eGFR 30–44) | 39 (32.5%) |
|  | Stage 4 (eGFR 15–29) | 28 (23.3%) |
|  | Stage 5 (eGFR <15 or on dialysis) | 16 (13.3%) |
| Progression Status | Progressors | 58 (48.3%) |
|  | Non-progressors | 62 (51.7%) |

**Table 2.**
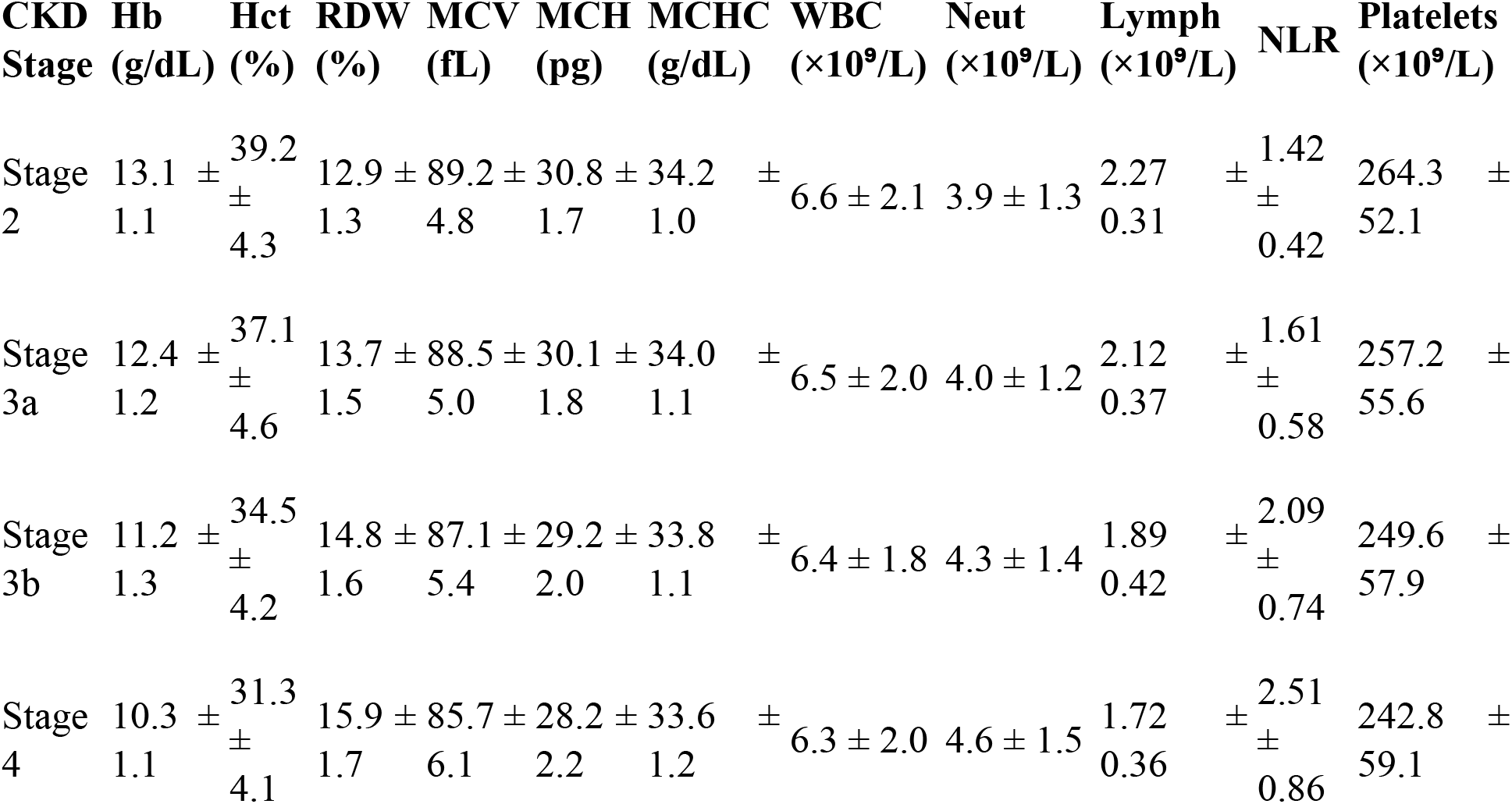
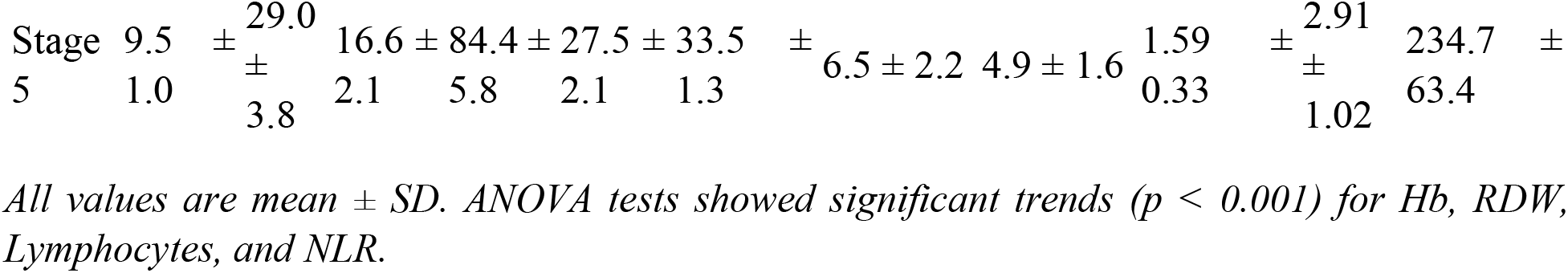
Hematological Indicators Across CKD Stages.

Comparison was done of hematological markers across varying CKD stages to chart the trend with progression of the disease by one-way ANOVA with a significant trend declared at p < 0.001. A marked worsening was noted in the key hematological indices as the CKD stage worsened. However, while a decrease in hemoglobin and lymphocytes and an increase in RDW and NLR were noted from stage 2 to stage 5 (p for trend <0.001).

### Comparison of Hematological Indicators by CKD Progression Status

Hematological parameters were compared by independent samples t-test between CKD progressors and non-progressors, with factors with P <0.05 being considered significant. Differences were noted in several hematological parameters between progressors and non-progressors. Those with progression had: Lower values of Hemoglobin, Hematocrit, MCV, MCH, and lymphocyte count; higher values of RDW and neutrophil-to-lymphocyte ratio (NLR).as shown in table 3 and figure 1. No significant differences were found for WBC total count, neutrophils, platelets, or MCHC.

**Table 3.** Hematological Parameters in Progressors vs. Non-Progressors.

| Parameter | Progressors (n = 58) | Non-progressors (n = 62) | p-value |
| --- | --- | --- | --- |
| Hemoglobin (g/dL) | 10.4 ± 1.2 | 12.2 ± 1.3 | <0.001 ** |
| Hematocrit (%) | 31.6 ± 4.5 | 36.4 ± 5.1 | <0.001 ** |
| Red Cell Distribution Width (RDW, %) | 15.6 ± 1.9 | 13.9 ± 1.7 | 0.002 ** |
| Mean Corpuscular Volume (MCV, fL) | 85.2 ± 6.1 | 88.4 ± 5.3 | 0.011 * |
| Mean Corpuscular Hemoglobin (MCH, pg) | 28.4 ± 2.2 | 30.2 ± 2.0 | 0.008 ** |
| Mean Corpuscular Hemoglobin Concentration (MCHC, g/dL) | 33.7 ± 1.1 | 33.9 ± 1.2 | 0.318 |
| White Blood Cells ($\times 10^9/L$) | $6.47 \pm 1.91$ | $6.72 \pm 2.11$ | 0.512 |
| Neutrophils ($\times 10^9/L$) | $4.18 \pm 1.35$ | $3.76 \pm 1.29$ | 0.064 |
| **Lymphocytes ($\times 10^9/L$)** | $1.83 \pm 0.41$ | $2.11 \pm 0.45$ | 0.003 ** |
| **Neutrophil-to-Lymphocyte Ratio (NLR)** | $2.43 \pm 0.92$ | $1.78 \pm 0.68$ | <0.001 ** |
| Platelets ($\times 10^9/L$) | $248.5 \pm 58.3$ | $252.8 \pm 61.2$ | 0.672 |

\*Values presented as mean ± SD. $p < 0.01$ , $p < 0.05$ .

**Figure 1.**
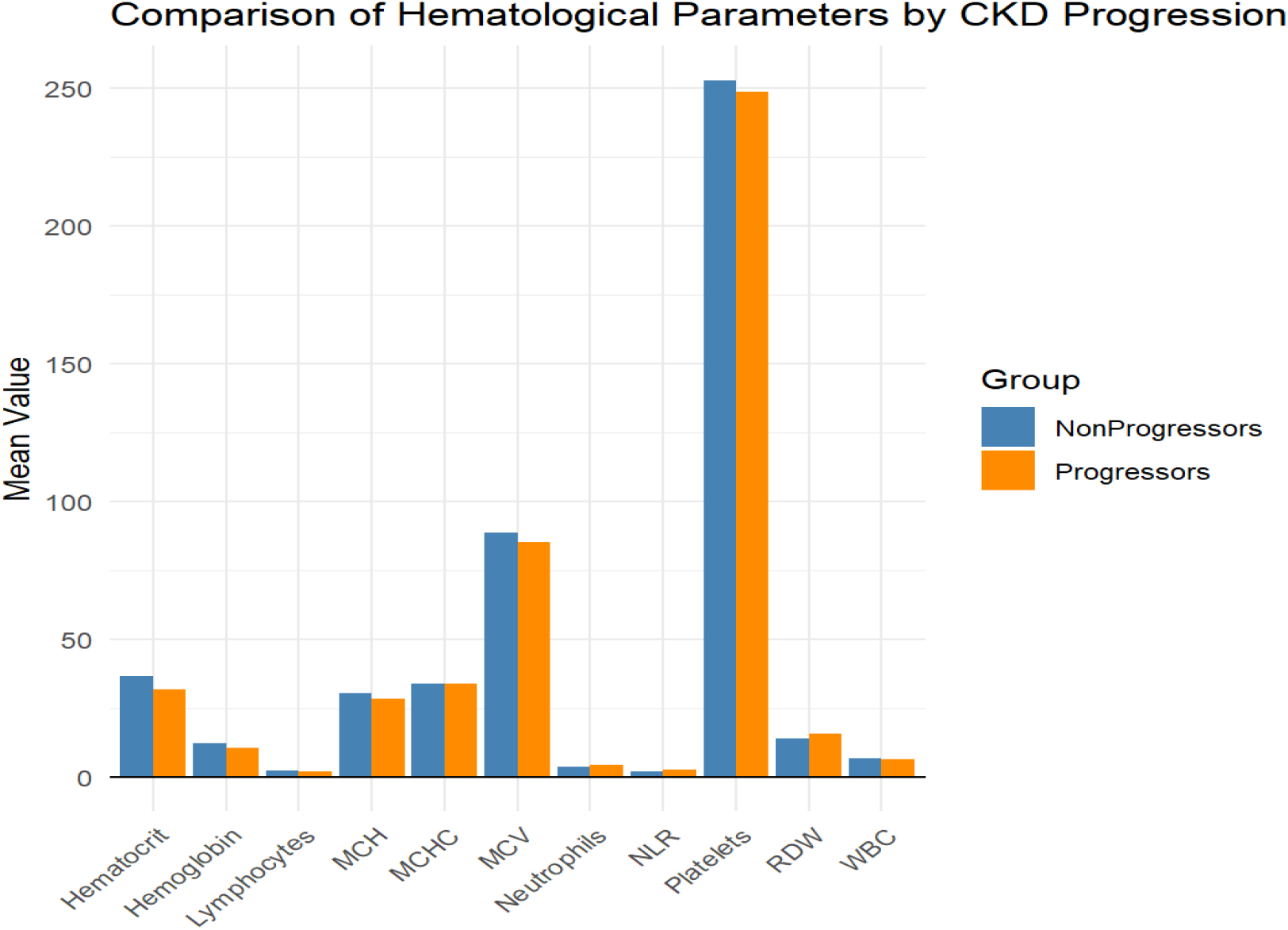
Hematological Parameters in Progressors vs. Non-Progressors.

### Multivariate Logistic Regression: Predictors of CKD Progression

Multivariate logistic regression was performed to identify independent hematological predictors for CKD progression. Adjusted odds ratios (aORs), 95% confidence intervals (CIs), and p-values for each predictor were described. Independent predictors of CKD progression from the multivariate logistic regression model were: Lower Hemoglobin (aOR = 0.57, 95% CI: 0.43–0.75, p < 0.001), Higher RDW (aOR = 1.51, 95% CI: 1.14–2.02, p = 0.005), Higher NLR (aOR = 1.89, 95% CI: 1.28–2.79, p = 0.002), and Lower Lymphocyte Count (aOR = 0.46, 95% CI: 0.21–0.99, p = 0.048) as illustrated in table 4 and figure 2. Other variables such as MCH, WBC, and platelet count were not significantly associated with progression.

**Table 4.** Multivariate Logistic Regression for Predicting CKD Progression.

| Predictor | Adjusted OR | 95% CI | p-value |
| --- | --- | --- | --- |
| Hemoglobin | 0.57 | 0.43 – 0.75 | <0.001 ** |
| RDW | 1.51 | 1.14 – 2.02 | 0.005 ** |
| NLR | 1.89 | 1.28 – 2.79 | 0.002 ** |
| Lymphocyte Count | 0.46 | 0.21 – 0.99 | 0.048 * |
| MCH | 0.91 | 0.78 – 1.07 | 0.270 |
| Platelets | 1.00 | 0.99 – 1.01 | 0.675 |

**figure 2.**
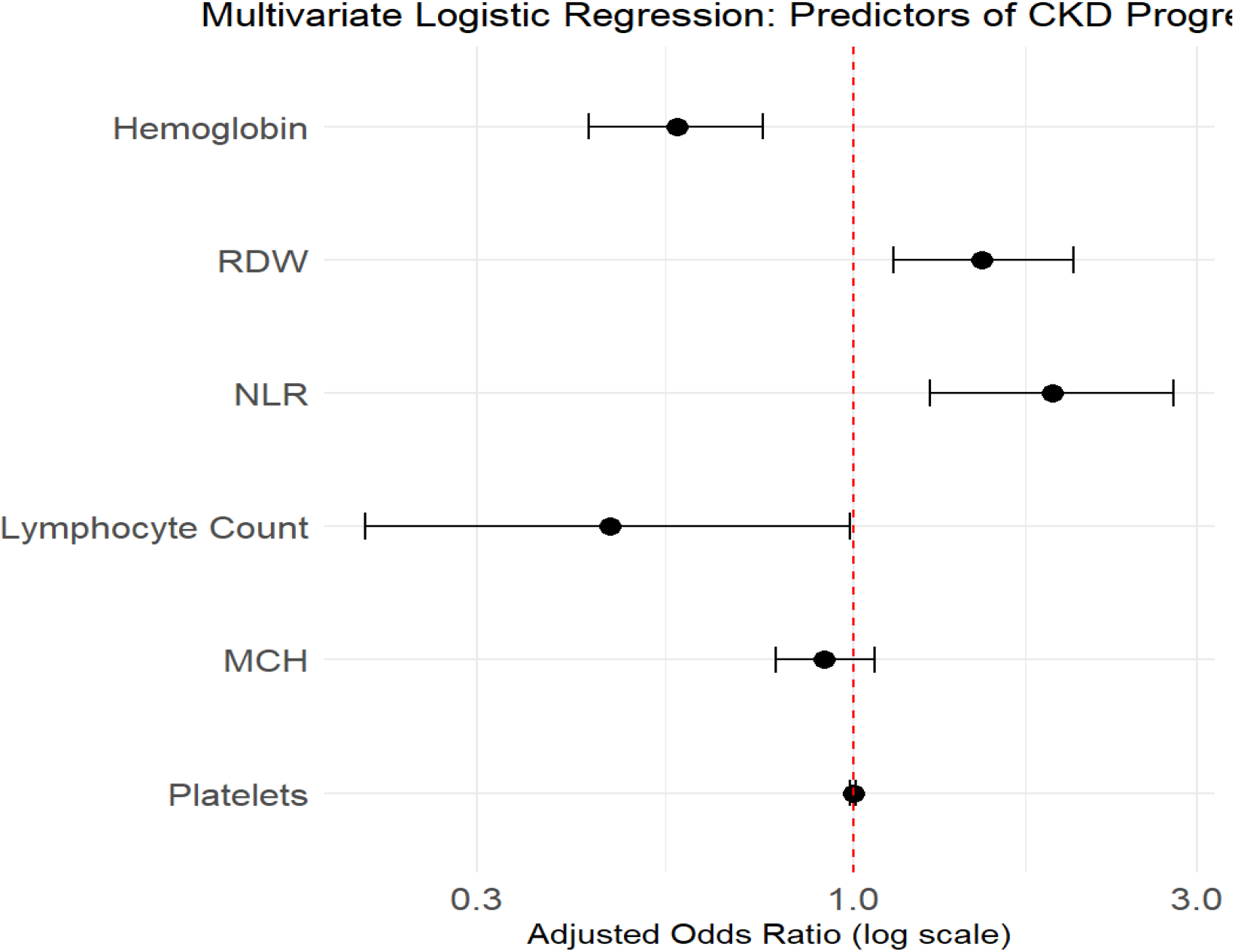
Multivariate Logistic Regression for Predicting CKD Progression.

## Discussion

This retrospective study sought to assess the prognostic significance of various hematological parameters in predicting CKD progression in patients at Kisii Teaching and Referral Hospital. Our study found hemoglobin (Hb), red cell distribution width (RDW), neutrophil-to-lymphocyte ratio (NLR), and lymphocyte count to be significant independent predictors of CKD progression, thereby supporting and adding to existing literature.

The major decline in Hb and hematocrit with advancing CKD stages in our study is consistent with the well-established anemia of CKD. Anemia occurs mainly due to decreased synthesis of erythropoietin by diseased kidneys, iron deficiency, and an inflammatory environment that inhibits erythropoiesis [9]. These factors were corroborated by previous reports from well-designed big cohort studies before ours (9), in which low Hb levels were seen as associated with faster renal decline and greater cardiovascular morbidity in populations with CKD. Our corroboration that Hb reduction independently predicts progression places the anemia centrally not only as a complication but also as a reason for the worsening of kidney function. Tissue hypoxia brought on by anemia could continue to escalate renal fibrosis, a vital route in the progression of CKD [10].

Raised RDW, indicating a variation in size of red cells in circulation, has lately become a point of attention as a prognostic factor in CKD. The association between increased RDW and CKD progression in our cohort backs previous claims by (11), who showed RDW to be an independent predictor of mortality and renal outcomes in CKD. It is sensibly deduced that RDW signifies chronic inflammation, oxidative stress, and nutritional deficiencies—factors that cause endothelial dysfunction and progressive nephron loss. Our data underscore RDW’s utility as a readily available, inexpensive biomarker that captures these complex pathophysiological processes.

With time, Neutrophil-to-Lymphocyte Ratio (NLR) is gaining increasing recognition as a sensitive systemic inflammation marker. We observed a higher NLR in progressors, in agreement with the studies by (12) who reported that elevated NLR portended poor renal and cardiovascular outcomes. Chronic inflammation is the key driver of renal fibrosis, and inflammatory cytokines mediate tissue damage while recruiting profibrotic cells that lay down scar tissue [13]. The decreased lymphocyte count in progressors also attests to a condition of immune dysregulation and might further signal defective immune surveillance or an ongoing chronic stress response, as also found in a CKD cohort by [14].

Mean corpuscular volume (MCV), mean corpuscular hemoglobin concentration (MCHC), total white blood cell count, and platelets were not independently associated with progression in our multivariate model. The conclusion may be that whilst these indices indicate any underlying derangement in the hematological status of the subject, they do not specifically predict worsening of CKD, a finding corroborated by previous smaller studies [15].

Our findings underscore the prognostic utility of routine hematological parameters in the management of CKD. Since these are widely available and inexpensive,, incorporating Hb, RDW, NLR, and lymphocyte count in risk stratification protocols could facilitate early identification of high-risk patients and optimize timely interventions. Larger prospective cohorts and mechanistic studies are required in future to confirm these markers and to investigate their potential role in directing therapies such as anti-inflammatory treatment or erythropoiesis-stimulating agents.

## Limitations

The retrospective design limits the prospects of inferring causality, and this being a single-center study may affect the generalizability of the results. Data were not available on other confounding variables such as nutritional status, iron studies, and comorbidities, which could have influenced hematological parameters. Nevertheless, this study stands out for its comprehensive assessment of a multitude of hematological indexes evaluated concerning CKD progression and with robust statistical associations, insights that can be of practical use in resource-limited settings.

## Conclusion

In brief, this study reveals that routine hematological parameters, namely low hemoglobin, increased RDW, high NLR, and low lymphocyte counts, are significantly correlated with CKD progression. This finding establishes the potential application of cheap and readily available blood indices as prognostic markers in the care of CKD, especially in resource-limited settings. If monitored regularly, this would facilitate early identification of high-risk patients and allow prompt intervention targeted toward slowing disease progression.

## Data Availability

All relevant data are within the manuscript and its Supporting Information files.

## Declarations

### Data availability

The data of the findings of this study are all shared on this article

### Consent for publication

All authors has given their consent for publication of this article

### Ethics approval and consent to participate

The study observed ethics according to Helsinki declaration and all participants signed voluntary written informed consent form.

## Authors’ contributions

**Author:** Collince Ogolla, Msc. **Contribution:** Conceptualization, development of the manuscript and review, data collection

**Author:** Dr. Lucy Karani, PhD **Contribution:** Review and supervision

**Author:** Dr. Stanslaus Musyoki, PhD **Contribution:** Review and Supervison

**Author:** Phidelis Maruti, Msc. **Contribution:** Review and Supervision

## Acknowledgment

N/A

## Authors’ Information

N/A

